# VTE incidence shortens survival in IDH-WT glioblastoma

**DOI:** 10.1101/2025.01.08.25319908

**Authors:** Anthony R. Sloan, Alan J. Gordillo, Austin Kennemer, Alok A. Khorana, Craig Horbinski, David C. Kaelber, Scott J. Cameron, Justin D. Lathia

## Abstract

**Background:** Venous thromboembolisms (VTE’s) are the second leading cause of death in cancer patients. While previous analyses have demonstrated VTE rates are greater in GBM patients using smaller patient cohorts in high-grade glioma, since the release of the update 5^th^ edition of the World Health Organization (WHO) classification a systematic analysis in a large-scale cohort of patients with IDH-wildtype GBM with clinical outcomes is lacking.

**Methods:** This study utilizes the online database, TriNetx, to build patient cohorts for outcomes analysis. TriNetX is a database comprised of over 50 healthcare organization patient information that is quarriable by CPT, ICD, RxNorm, and other proprietary codes. Patient cohort demographics were used for propensity score matching. Risk ratios, odds ratios, hazard ratios, and Kaplan Meier curves were utilized for primary outcomes including survival and time-to-event analyses.

**Results:** 24% of patients with GBM experienced at least 1 VTE or PE after their diagnosis. Compared to a population of patients with no cancer history with an index event of an inpatient visit, patients with GBM were at 20.4 (12.23-34.17) and 5.96 (3.85-9.23) times higher risk of experiencing a VTE/PE at 1- and 5-year follow-up, respectively. Sex differences were not seen between VTE/PE rates and survival after VTE/PE at 1- and 5-year follow-up (p>0.05). Lastly, patients with GBM and a VTE/PE after diagnosis experienced worse survival at 1- and 5-year follow-up compared to those without a VTE/PE (p<0.0001 and p = 0.0014, respectively).

**Conclusions:** Patients with GBM experience increased risks of thrombotic events after diagnosis. These risks are not sex-dependent but do affect overall survival.

---

Cancer-related thrombosis, specifically venous thromboembolism (VTE) consisting of either one of—or both—deep vein thrombosis (DVT) or pulmonary embolism (PE), is a common co-morbid condition in patients with cancer^1^. Isocitrate dehydrogenase (IDH) wild-type glioblastoma (GBM) is an underappreciated disease with increased VTE risk. GBM is highly-vascularized and produces angiogenic factors, with 92% of GBM resections showing histological evidence of intravascular thrombosis; this is greater than any other CNS tumor^2^. Moreover, thrombosis-related signaling due to aberrant platelet activation has been reported in GBM^3, 4^, underscoring these mechanisms as potential drivers of GBM growth.

While previous analyses have demonstrated VTE rates are greater in GBM patients using smaller patient cohorts in high-grade glioma^5-8^, since the release of the update 5^th^ edition of the World Health Organization (WHO) classification a systematic analysis in a large-scale cohort of patients with IDH-wildtype GBM with clinical outcomes is lacking. To address this unanswered question, we leveraged the Research USA Minimal Date Shift network in the TriNetX platform composed of aggregated, de-identified electronic health record data of 93 million from various healthcare organizations (HCOs) nationwide. Inclusion criteria for general GBM comparisons included diagnosis of IDH-wildtype GBM. Healthy patient comparison inclusion criteria were an in-patient hospital stay for non-acute causes with no neoplasm diagnosis. Index events were queried such that their occurrence must fall within the years 2000-2023 for 1-year follow-up data and 2000-2019 for 5-year follow-up data. Queries were assessed on 10/2024. VTE was selected as any instance of the ICD-10-CM codes I82 (other venous embolism and thrombosis) and I26 (PE). Log-Rank analysis was performed using sex-specific Kaplan Meier survival curve models adjusted for age at index event, sex, and age at diagnosis.

The study population comprised 1535 healthy patients,1496 patients diagnosed with IDH-wildtype GBM with a VTE, and a cohort of 1,138 patients with IDH-wildtype GBM with no VTE. Propensity-matched clinical cases were evaluated based on age at index, race, sex, ethnicity, metabolic disorders, hypertension, ischemic heart disease, and diabetes. After propensity-matching, each cohort was well-balanced with standardized differences with p<0.01. Patients with GBM had a 24% incidence of VTE compared to 2.3% incidence for inpatient hospitalizations for all causes (OR=13.3, 95% CI 9.3-19.0). At 1 year follow up, GBM patients had a 19.1% rate of VTE compared to 1.394% in the healthy non-neoplastic population (OR=25.3, 95%CI = 15.0-42.8). At 5 years follow up, GBM patients had a 21.9% rate of VTE compared to 3.2% in the healthy non-neoplastic population (OR = 7.5, 95% CI = 4.7-12.0).

At both the 1 and 5 years following GBM diagnosis, after propensity score matching as described above, patients with a VTE had a significantly higher risk for all-cause mortality compared to patients without a clinical diagnosis of VTE (**Fig. 1**, HR=0.5, 95% CI = 0.4-0.7; HR=0.6, 95% CI = 0.5-0.94). Of note, there was no significant difference in mortality risk for males and females 1 year following GBM diagnosis or 5 years following GBM diagnosis.

**Fig. 1.**
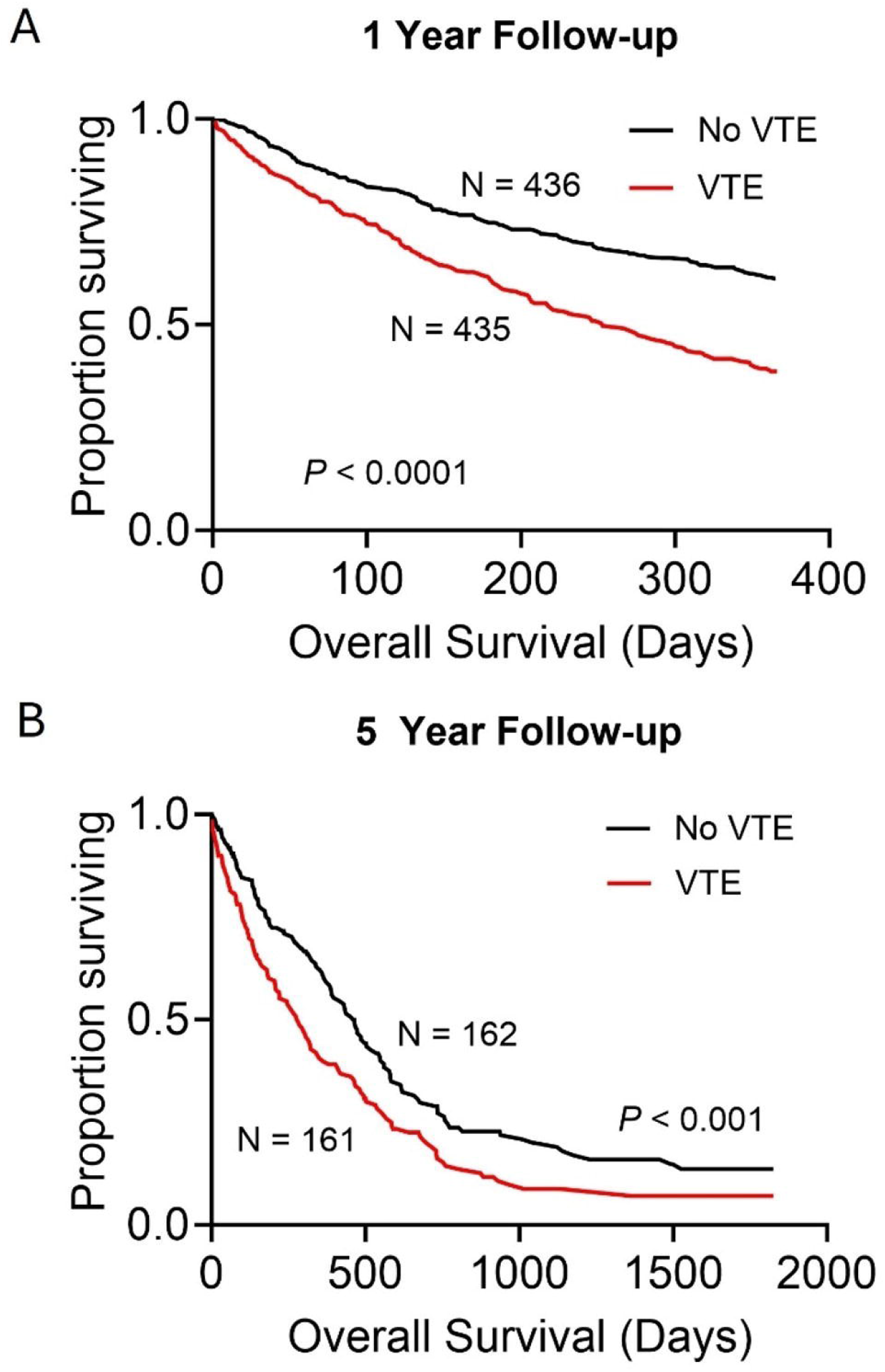
(A) 1-year survival of IDH-wt GBM patients diagnosed with a VTE and those with no VTE. (B) 5-year survival of IDH-wt GBM patients diagnosed with a VTE and those with no VTE.

The results of this analysis confirm greater than 20% of IDH-WT GBM patients develop VTE. Separate analysis with unique cohorts was done to compare methylation of the O^6^-methylguanine-DNA methyltransferase (MGMT) promoter status, sex, and IDH status, which were not predictive of VTE incidence.. We acknowledge a prior study using LASSO did indicate a protective effect of IDH and MGMT methylation status^5^. A limitation of this study—or any study relying on administrative databases—is the possibility of incorrect coding for diagnoses at the time of clinical care.

A limitation to this analysis using TriNetX platform is lack of information regarding cause of mortality, which future studies should address. However, a previous report has demonstrated that although VTE incidence is a leading cause of death in patients with GBM, only 1% of GBM-related deaths are a consequence of VTE, whereas a 90% of patients with GBM and a VTE die because of tumor progression^6^. Mechanistic data are needed to clearly understand how platelets and the coagulation cascade are primed toward a thrombotic state to identify how these mechanisms uniquely contribute to GBM disease progression, identifying therapeutic targets.

## Data Availability

All data produced in the present study are available upon reasonable request to the authors

## Authorship

ARS, JDL conceptualized the project; ARS, AJG, AK completed the analysis; All authors participated in the writing and review of the manuscript.

## Acknowledgements

The authors thank Dr. Erin Mulkearns-Hubert for editorial assistance and Dr. Reza Khatib for his inspiration and support.

